# Genetic factors affecting immune phenotypes in type 1 diabetes

**DOI:** 10.1101/2021.12.06.21264056

**Authors:** Xiaojing Chu, Anna W.M. Janssen, Hans Koenen, Linzhung Chang, Xuehui He, Irma Joosten, Rinke Stienstra, Yunus Kuijpers, Cisca Wijmenga, Cheng-Jian Xu, Mihai Netea, Cees J. Tack, Yang Li

**Affiliations:** Department of Genetics, University of Groningen, University Medical Center Groningen; Centre for Individualised Infection Medicine, CiiM, a joint venture between the Hannover Medical School and the Helmholtz Centre for Infection Research, Hannover, Germany; TWINCORE, Centre for Experimental and Clinical Infection Research, a joint venture between the Hannover Medical School and the Helmholtz Centre for Infection Research, Hannover, Germany; Department of Internal Medicine, Radboud University Medical Center, Nijmegen, the Netherlands.; Department of Laboratory Medicine, Laboratory Medical Immunology, Radboud University Medical Center, Nijmegen, the Netherlands.; Division of Human Nutrition and Health, Wageningen University, the Netherlands.; Department for Genomics & Immunoregulation, Life and Medical Sciences Institute (LIMES), University of Bonn, Bonn, Germany.

**Keywords:** functional genomics, type 1 diabetes, immune function

## Abstract

Large inter-individual variability in immunological cell composition and function determines immune responses in general and susceptibility to immune-mediated diseases in particular. While much has been learned about the genetic variants relevant for type 1 diabetes, the pathophysiological mechanisms through which these variations exert their effects are unknown. In this study, we characterize the genetic factors influencing immune responses in patients with type 1 diabetes. Genetic variants that determine susceptibility to T1D significantly affect T cell composition. Specifically, the CCR5+ regulatory T cells associate with T1D through the CCR region, suggesting a shared genetic regulation. Genome-wide quantitative trait loci (QTL) mapping analysis of immune traits revealed 15 genetic loci that influence immune responses in T1D. Among them, 12 have never been reported in healthy population studies, implying a disease-specific genetic regulation. Altogether this study provides new insights into the genetic factors that affect immunological responses in T1D.

## Introduction

T1D is a common, chronic, autoimmune disease, characterized by destruction of insulin- producing beta-cells in the pancreas that results in lifelong dependence on exogenous insulin and is associated with a high morbidity and mortality *(Atkinson et al., 2014)*. The causes and immunological pathways responsible for the development of T1D are still incompletely understood, which hampers the efforts to identify an etiopathogenetic treatment.

Many studies have highlighted the role of environmental, genetical, and immunological factors in the pathogenesis of T1D*(Pociot and Lernmark, 2016; Rewers and Ludvigsson, 2016)*.

Environmental factors such as overweight, infections, microbiome composition and dietary deficiencies have been reported as risk factors for T1D*(Rewers and Ludvigsson, 2016)*. In turn, the immunological pathogenesis*(Cabrera et al., 2016)* of T1D includes innate inflammation and adaptive immunity, such as enhanced T cell responses*(Hundhausen et al., 2016)*. Large genome- wide association studies (GWAS) performed in the last two decades have underscored the contribution of genetic polymorphisms for the susceptibility to T1D: ∼60 genomic loci associated with T1D risk have been identified (Barrett et al., 2009; Bradfield et al., 2011; Cooper et al., 2008; Grant et al., 2009; Huang et al., 2012; Onengut-Gumuscu et al., 2015; Ram et al., 2016). While these loci show significant enrichment in specific immune-related biological pathways such as cytokine signaling and T cell activation*(Barrett et al., 2009; Cooper et al., 2008)*, the functional consequences of many of these loci and genetic variants are still unknown. We thus lack the information that could link the genetic susceptibility factors to the immunological pathways potentially important for T1D pathogenesis. The genetically-regulated inflammatory response signature in T1D may also be relevant for the inflammatory response in general and may become modified by the chronic hyperglycemic state.

In the present study we aimed to comprehensively describe the immunopathological consequences of the genetic variants linked to T1D susceptibility, using a high-throughput functional genomics approach. As a part of the Human Functional Genomics Project (HFGP)*(Netea et al., 2016)*, we carried out deep immunophenotyping in peripheral blood samples from a cohort of 243 T1D patients (300DM), covering cell subpopulation composition and cytokine production upon stimulations, as proxies of immunological function. Part of the results were compared to those obtained in a population-based cohort of 500 healthy individuals (500FG), that successfully characterized the impact of genetic factors *(Aguirre-Gamboa et al., 2016; Li et al., 2016)* on immune responses in healthy individuals. Here, we systematically evaluate the genetic regulation on the immune phenotypes in T1D, and show how genetic variation affects immune cell traits and cytokine production in response to stimulations. In total, we identified 15 genome wide significant genomic loci (P value < 5 × 10^-8^) associated with immune phenotypes in the 300DM cohort. Among them, 12 loci have never been reported in any healthy population study. These data provide a deeper understanding of the immune mechanisms involved in the pathophysiology of T1D and affecting the general inflammatory response and may open avenues towards the development of novel diagnostics and potentially immunotherapies.

## Results

### Interrelationship between immune cell counts and cytokine production in T1D

We collected blood samples from 243 T1D patients (300DM cohort, Figure 1—figure supplement 1), following the same methodology as previously described (Aguirre-Gamboa et al., 2016; ter Horst et al., 2016; Li et al., 2016). The baseline characteristics of the 300DM and a cohort of healthy individuals (500FG) are shown in Supplementary file 1. Their median age was 53.5 years (range 20-85), and they had a median diabetes duration of 28 years (range 1-71 yrs).

**Figure 1.**
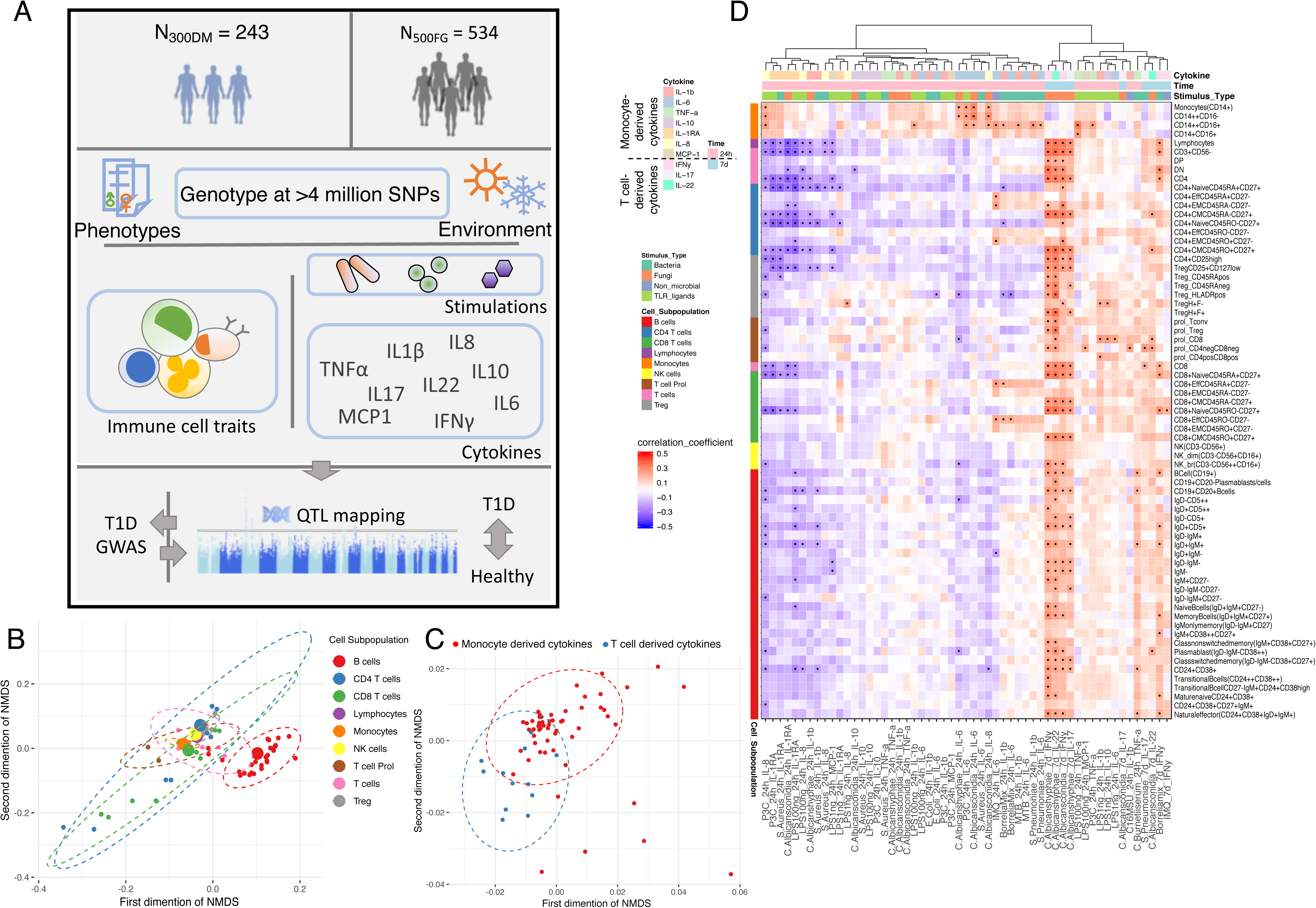
Overview of data and experimental design. **A**, Schematic study design. **B**,**C**, Cell (**B**) and cytokine (**C**) type relationship visualized by non-metric multidimensional scale (NMDS) analysis in 300DM, where B cell traits (red) are clustered separately from T cell and monocyte traits (B) and cytokines with same cellular origins are clustered together (C) In panel **B**, Small dots indicate the proportion of subpopulations and large dots indicate counts of their parental cell types. Circles represent the calculated centroid of the grouped cell and cytokine types at confidence level 0.95. **D**, Heatmap of correlation coefficients between immune cell counts (y-axis) and cytokine production in response to stimulations (x-axis) in T1D patients. Significant correlations (FDR <0.05) are labeled by dots, with the color indicating correlation coefficients. Positive correlations (red) are observed between monocyte traits and monocyte-derived cytokines (top left), and between adaptive immune cell counts and T cell-derived cytokines (bottom right). Negative correlations (blue) are observed between adaptive immune cell counts and monocyte-derived cytokines.

Hence, the cohort generally consisted of middle-aged people with long standing type 1 diabetes. We measured 72 types of immune cells covering both lymphocytes and monocyte lineages and 10/6 (300DM/500FG) different cytokines released in response to stimulation with four types of human pathogens in both cohorts (Figure 1A, Figure 1—figure supplement 2).

The nonmetric multidimensional scaling plot illustrates the interrelationship among immune cells’ abundance (Figure 1B), and cytokine production levels (Figure 1C) in T1D patients. We observed a separation of the B cell subpopulations cluster from the T cell, monocyte, and NK cell population clusters. This observation suggests that T cells, monocytes, and NK cells have more interplay than B cells at baseline, which is consistent with our previous finding in a healthy cohort (500FG(Aguirre-Gamboa et al., 2016)). Furthermore, cytokine features are clustered based on their cellular origins, with a partially overlapping cluster between monocyte-derived cytokines and T cell-derived cytokines, Figure 1C). This may suggest activation of a co-regulatory network of monocyte-derived and T cell-derived cytokine production capacity in T1D.

To obtain a comprehensive interaction map between immune cells and cytokines, we correlated each of the immune cell counts with each of the cytokine production profiles (IL-1β, IL-6, TNF- α, IL-10, IL-1RA, IL-8, MCP-1, IFNγ, IL-17, and IL-22) in response to 21 stimulations.

Cytokine levels were hierarchically clustered based on correlation coefficients with immune cell counts (Figure 1D). In line with the functional relationship between immune cells and cytokines, we observed positive correlations between monocyte lineages and monocyte-derived cytokines (IL-1β, IL-6, TNF-α, IL-10, IL-1RA, IL-8, and MCP-1), as well as between T cell subsets and T cell-derived cytokines (IFNy, IL-17, and IL-22) (indicated by red color in Figure 1D). Besides, we found a negative correlation between monocyte-derived cytokines production in response to four distinct types of stimulations (bacteria, fungi, non-microbial and TLR ligands) and lymphocyte counts (blue, Figure 1D). We observed similar correlation patterns in 500FG (Figure 1—figure supplement 3). This correlation matches the findings that a high abundance at baseline of adaptive immune cells is associated with a lower production of monocytes derived cytokines after stimulation*(Dong Kim et al., 2007)*, which is not altered by T1D status. Overall, the inter- relationships between immune cells and cytokines in response to stimulations are rather similar in T1D as compared to healthy individuals.

### Impact of T1D GWAS SNPs on immune phenotypes in T1D patients

Considering that T1D is a multifactorial disease with a genetic component, we tested whether the known risk variants of T1D affect immune phenotypes and function. We firstly checked SNPs within HLA locus in our association studies on cell proportion and cytokine production level.

Consistent with our previous findings in 500FG, we did not observe any significant associations of HLA allelic variants in 300DM.

Non-HLA genetic loci from published GWAS studies of European background were acquired from the GWAS-catalog (Nov 2019)*(Buniello et al., 2019)*. Among them, genetic variants in 63 independent T1D loci were present in our data, and we found that 13 out of these 63, indeed associated with susceptibility to T1D with nominal significance (P value < 0.05) (Supplementary file 2).

Next, we investigated whether these genetic risk loci for T1D affect immune parameters and function. The quantile-quantile (Q-Q) plot of the association of the 63 T1D GWAS loci with different cell types and cytokines illustrates an inflated deviation from an expected uniform distribution (Figure 2A, Figure 2—figure supplement 1). We further tested whether this deviation can be explained by chance by comparing the association of immune traits with T1D GWAS SNPs with that of 1000 randomly selected independent SNPs (Figure 2B, Methods). The p value shows that the T1D GWAS SNPs are enriched in association with T-cell traits in the T1D cohort (P value = 0.007).

**Figure 2.**
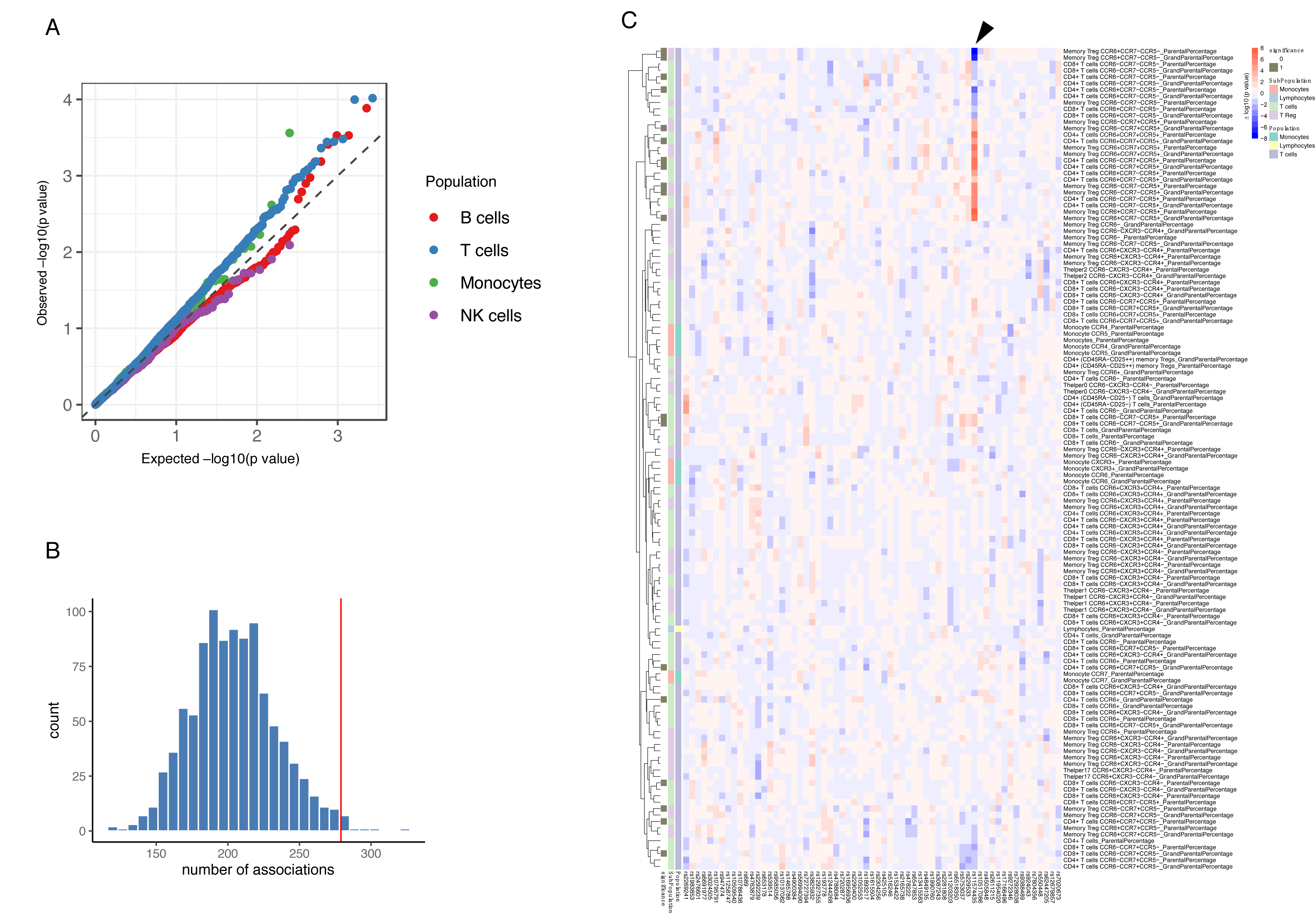
Impact of T1D GWAS SNPs on immune phenotypes. **A**, QQ plots of QTL profiles of 62 T1D GWAS loci grouped by cell populations. The distribution of P values of associations with T cells traits (blue) show a significant deviation from an expected uniform distribution (dashed line). **B**, Histogram showing number of associations observed (red line) and those in permutations (blue bars). **C**, Heatmap of QTL profiles of cell proportion carrying certain chemokine receptors across 62 T1D GWAS loci, colored by -log10(p values) and effect direction of T1D risk allele. Arrowhead indicates a T1D risk allele rs11574435-T.

A pair-wise association analysis between T1D GWAS loci and immune phenotypes shows that 261 out of 269 immune cell phenotypes and 53 out of 55 cytokine-stimulation pairs are suggestively associated with at least one T1D GWAS locus (P value < 0.05, Supplementary files 3, 4). We further applied a permutation-based approach to test whether immune phenotypes were significantly influenced by the accumulative effects of these 63 GWAS loci (Methods).

Compared to random sets of independent SNPs, the 63 T1D GWAS loci explain significantly more variance of 27 cell sub proportions and 15 cytokine production traits (P value < 0.05, Figure 2C, Figure 2—figure supplement 2A, B). As shown in the heatmap (Figure 2C, arrowhead), a T1D risk allele rs11574435-T, in strong LD (r^2^ = 0.95) with the T1D GWAS SNP rs113010081*(Onengut-Gumuscu et al., 2015)*, is associated with higher percentage of many CCR5+ CD4+ T-cell traits and lower percentage of CCR5- CD4+ T-cell traits. Chemokine signaling pathways regulate the migration of cells from the circulation (PBMCs) to the tissue (pancreases).

To further validate the importance of chemokine signaling mediated by *CCR5* in T1D, we illustrated the transcriptional changes on *CCR5* and its corresponding ligand genes using publicly available data from transcriptome analysis in PBMCs and pancreatic tissue from T1D patients and controls *(Planas et al., 2010; Yang et al., 2015)*(See also Methods), and found significant expression changes of *CCR5*, *CCL5*, and *CCL4* in T1D patients, suggesting the involvement of this chemokine ligand - chemokine receptor pathway (Figure 3A). Besides, another top SNP- rs35092096 within the *CCR* genes region has the strongest effects on many CCR5 Tregs proportions in T1D. For example, the minor allele T in rs35092096 associates with a higher ratio of CCR6+CCR7-CCR5+ Tregs/ CCR6+ T regs (Figure 3B, C). Together with the observed regulation from a GWAS locus within the *CCR* region on CCR6+CCR7-CCR5+ Tregs proportion, we tested whether CCR6+CCR7-CCR5+ Tregs and T1D shared the same causal variants/genomic regions by integrating the cell proportion QTL of CCR6+CCR7-CCR5+ Tregs and the latest T1D GWAS profile using colocalization analysis*(Giambartolomei et al., 2014)*. The result strongly supports that CCR6+CCR7-CCR5+ Tregs share the same regulatory genomic region with T1D, although the causal SNP might be different (Figure 3D, H3 = 0.95). Altogether, these results support a role of the *CCR* region on Tregs function in the pathogenesis of T1D. Overall, we observed that T1D GWAS loci influence immune cell proportion and cytokine production capacity, again stressing the importance of T cell immunity in the genetic regulation of T1D.

**Figure 3.**
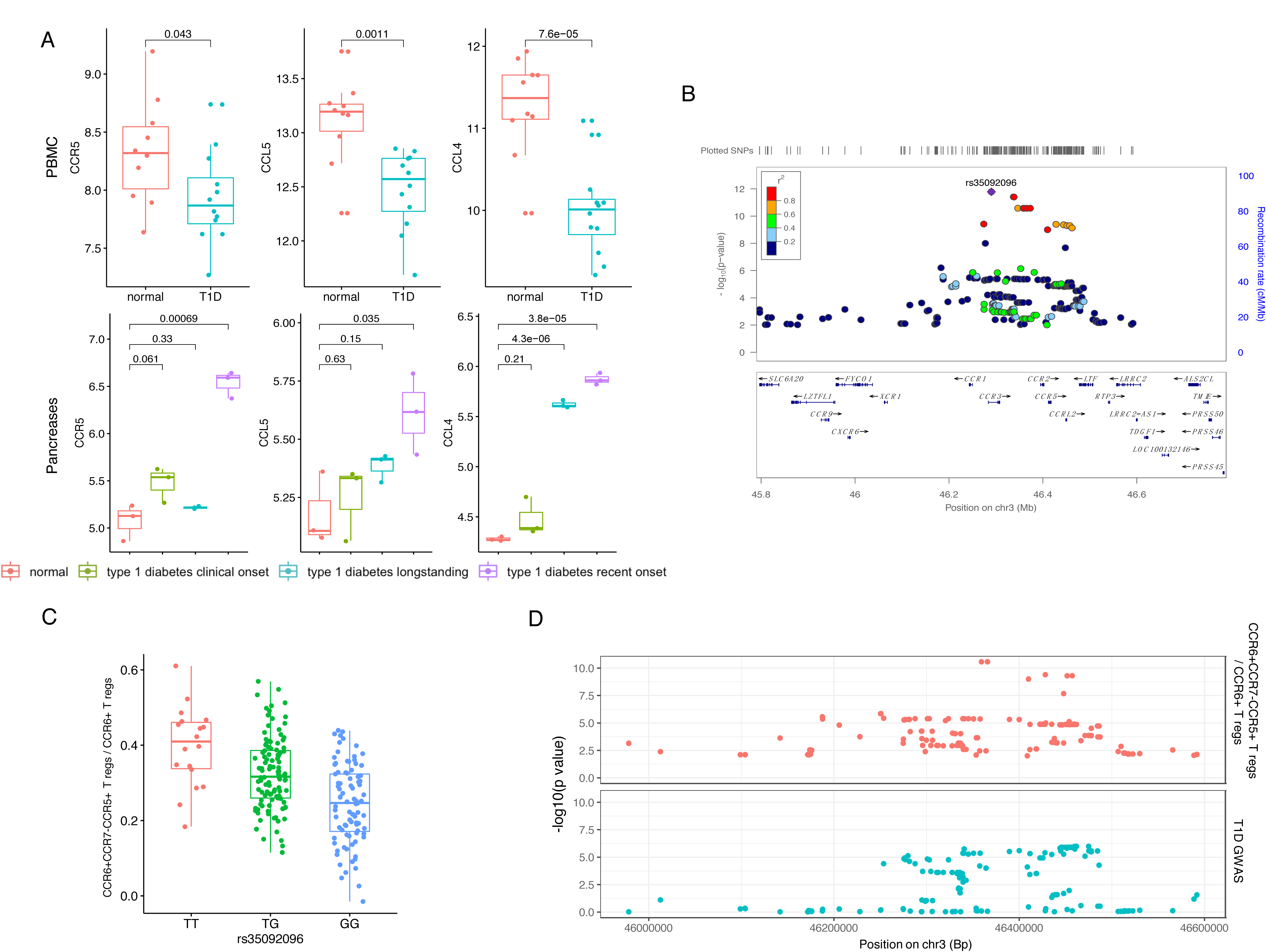
Impact of T1D GWAS SNPs on immune phenotypes. **A**, Expression of *CCR5*, *CCL5* and *CCL4* in PBMCs (top) and Pancreases (bottom) altered in T1D patients (blue, green and purple) compared to healthy controls (red). **B**, A locus zoom plot showing SNPs around rs35092096 located in CCR region are associated with CCR6+CCR7-CCR5+ T reg proportion. **C**, A boxplot showing CCR6+CCR7-CCR5+ T reg proportion differs in different rs35092096 genotypes (TT: red, TG: green and GG: blue). **D**, Two locus zoom plots indicating colocalization between CCR6+CCR7-CCR5+ T reg proportion QTL profiles (top, red) and T1D GWAS profile (bottom, blue) within CCR regions.

### Genetic regulation of immune phenotypes in T1D

To further explore potential genetic regulation of immune phenotypes on the whole-genome level, we performed quantitative trait loci (QTL) mapping in 300DM. We identified nine genome-wide significant QTLs (P value < 5x10^-8^) associated with immune cell proportion, including four associated with T cell subpopulations expressing specific chemokine receptors (e.g., rs35092096 and rs7614884) (Figure 4A, top and middle panels, Supplementary file 5). A pathway analysis of the cell proportion QTLs shows significant enrichment in chemokine and cytokine signaling-related biological pathways (FDR < 0.05, Figure 4—figure supplement 1A), highlighting the effects of immune signaling genes in cell proportion regulation.

**Figure 4.**
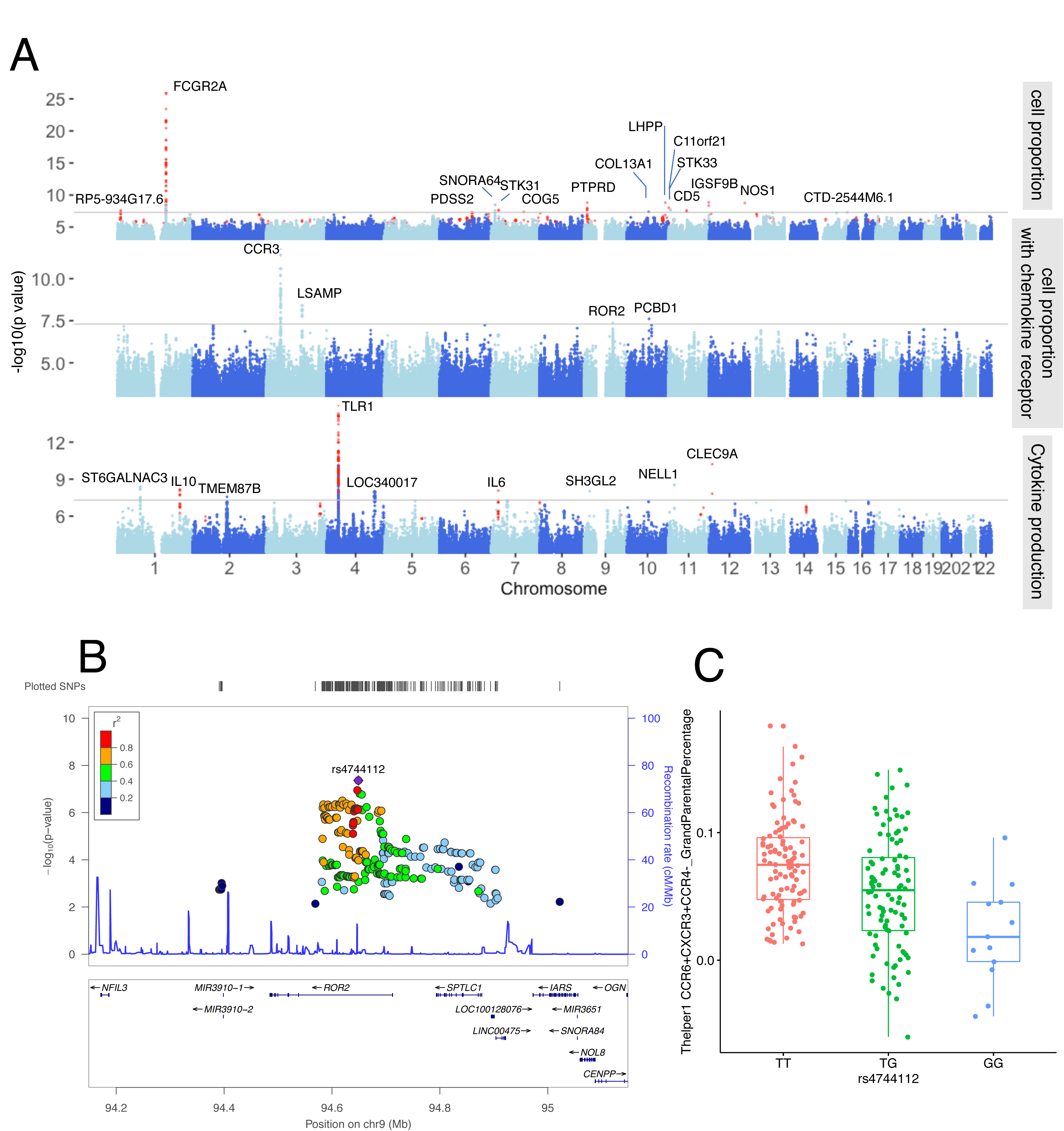
Genetic regulators on immune phenotypes. **A**, Three Manhattan plots showing genetic regulators of immune cell proportion (top), proportion of immune cells expressing CCR (middle) and cytokines production in response to stimulations (bottom). P values of SNPs identified in meta-analysis were colored in red. **B**, A locus zoom plot showing a T1D specific regulatory locus around rs4744112 that effects CCR6+CXCR3+CCR4- T helper1 proportion. **C**, A boxplot showing CCR6+CXCR3+CCR4- T helper1 proportion varies in different rs4744112 genotypes (TT: red, TG: green and GG: blue).

In parallel, we detected six significant genomic loci associated with cytokine production in response to stimulations by QTL mapping in 300DM (Figure 4A, bottom panel, Supplementary file 5). TLR-related pathways significantly enrich cytokine production QTLs (FDR < 0.05, Figure 4—figure supplement 1B). A missense variant, rs5743618, on the *TLR1* gene, affects IL-6 production in response to *S.pneumoniae* in PBMC. Previously, we observed the strong effect of the TLR locus on cytokine levels in controls*(Li et al., 2016)*. Our results support the reported function of TLRs in detecting pathogens (including bacterial) and triggering the release of inflammatory cytokines*(Fitzgerald and Kagan, 2020)*.

Next, to identify the consistent immune phenotype QTLs within T1D patients and healthy individuals, and to expand our understanding on genetic regulation of immune phenotypes, we applied a meta-analysis by integrating the obtained QTLs from the 300DM and 500FG cohorts. In total, we newly identified ten genetic loci that were associated with immune cell proportion (Figure 4A, top panel, colored in red) and three genetic loci that were associated with cytokine production (Figure 4A, bottom panel, colored in red), including a *CD5* locus significantly associated with IgD+ CD5++ proportion in B cells (Supplementary file 5). This may be important as recent studies report the relevance of IgD+ CD5++ B cells in T1D*(Saxena et al., 2017)* as well as within other autoimmune diseases such as Graves’ disease*(van der Weerd et al., 2013)*.

To summarize, we identified 28 genetic loci associated with either immune cell traits or cytokine production phenotypes. Among them, the genes within 11 loci (+- 250kb window) are druggable according to public drug database*(Finan et al., 2017)*, including *COL13A1*, *CCR* family genes, *ROR2*, *NOS1*, *IL-10*, *IL-6*, *STK33*, *NELL1, FCGR2A, CD5* and *TLR1* (Supplementary file 5).

Some of these genes are already therapeutic targets in other autoimmune diseases, such as the *STK33* inhibitor for rheumatoid arthritis treatment*(Rolf et al., 2015)*.

It is also worth mentioning that, as far as we know, 12 out of the 28 significant loci have never been reported in other healthy individual population cohorts according to records in PhenoScanner V2 (Kamat et al., 2019) (P value < 1 × 10^-5^, November, 2019), neither were they identified in the 500FG cohort (P value < 0.05). An example of a T1D specific locus is that rs4744112 located on chromosome 9 has an influence on CCR6+CXCR3+CCR4- Th1 like helper proportion (Figure 4B), with minor allele G leading to a decrease in these cells relative to major allele T (Figure 4C). rs4744112 is located within the transcriptional starting site (TSS) and enhancer regions*(Roadmap Epigenomics Consortium et al., 2015)* of *ROR2*.

### Functional Clues from the Associated Variants in T1D patients

In order to understand the mechanism behind the genetic regulations of immune response in T1D, we next explored the function of identified genetic factors of immune phenotypes. We noticed that SNPs within the 28 immune parameter QTLs are mostly located in the intergenic and intronic region (Figure 4—figure supplement 2), suggesting that identified genetic variants influence immune phenotypes through regulatory effects rather than through protein structure alteration. Moreover, seven out of 28 loci influence blood gene expression in public databases*(Carithers and Moore, 2015; Westra et al., 2013)* (Supplementary file 5) including rs7512140: FCGR2B/FCRLB, rs35092096: CCR1/CCR3, rs4744112: ROR2/SPTLC1, rs10840031: TRIM66, rs800139: C11orf21, rs1518110: IL10 and rs56350303: AC091814.2.

These indicate the potential functional genes behind the identified genetic loci.

## Discussion

The present study applies a high-throughput functional genomics approach to identify the association between genetic factors and the inflammatory phenotype in patients with T1D. The results confirm a correlation between baseline immune cell populations and ex vivo cytokine production in response to bacteria, fungi, non-microbial and TLR ligand stimulations. We provide evidence for a direct link between T1D GWAS loci and immune functionality, particularly on circulating T cell subpopulations. We show that T cell alteration is largely driven by T1D genetics, while B cells do not show a significant association with T1D GWAS loci. The association between the proportion of CCR5+ Tregs and T1D susceptibility through CCR genes suggests that T1D associated genetic variants contribute to immune function alteration through an accumulative effect. Finally, out of 28 genome-wide significant genetic loci regulating immune cell proportions and cytokine production, we identified 12 immune phenotype QTLs specific to 300DM. We also found 11 druggable genes as candidates for therapeutic intervention. Altogether, this study provides several novel insights into the genetic variability of immune traits in T1D.

Our finding of a correlation between baseline immune cells and cytokine production in response to bacteria, fungi, non-microbial and TLR ligand stimulations, suggests that steady-state immune cell abundance and alteration of immune cell proportion also affects immune response. This is in line with earlier findings on the regulation between immune cells and cytokine production*(Dong Kim et al., 2007)*. This finding has two important consequences: the immune response variability could be partly explained by variation in steady-state, and treatment of steady-state immune cells before stimulation could also influence immune response after stimulation.

Studies comparing T1D patients and healthy controls show that immune-related genes are physically located in T1D loci or differentially expressed. More studies highlight the importance of T cell immunity in T1D pathology*(Farh et al., 2015)*. In our study, we provide evidence for a direct link between T1D GWAS loci to immune functionality, particularly on circulating T cell subpopulations. We show that T cell alteration is largely genetically driven, while, interestingly, B cells do not show a significant association with T1D GWAS loci. It is thus tempting to speculate that the occurrence of auto-antibodies in T1D patients have limited pathophysiological relevance, but are merely markers of an autoimmune process mainly driven by T-cells as suggested before (Martin et al., 2001).

Importantly, we reveal that the effects of different GWAS loci on immune phenotypes vary. Despite the strong effects of the CCR locus (mean absolute effect sizes = 0.017 (cell proportion), 0.15 (cytokine production)), we observed much weaker individual effects from other T1D GWAS loci on immune cell proportion parameters (mean absolute effect size = 0.0060) and/or cytokine production (mean absolute effect size = 0.10) (Figure 2C and Figure 2—figure supplement 2). This finding indicates that T1D-associated genetic variants might alter immune function through a cumulative effect. Considering the complexity of the immune system, it may be an alternative explanation for our finding that T1D associated genetic variants that affect immune functions differ from genetic variants that affect immune cell proportion and cytokine production capacity.

Despite the limited sample size, we identified 28 genome-wide significant genetic loci regulating immune cell proportions and cytokine production in T1D patients and health. Among them, 12 immune phenotype QTLs are specifically found in 300DM, but not in healthy volunteers, suggesting a distinct regulatory mechanism of immune parameters and functions between disease and health. More importantly, in our study, we highlight 11 druggable genes as candidates for therapeutic intervention.

The data presented in our study are generated from PBMC. While these likely reflect overall immune function, some immune cell types may not be captured and all over the findings refer to changes in circulating factors that may not necessarily reflect changes occurring in relevant immune organs, such as pancreatic islets, gut or lymph nodes. Still, islet infiltrating immune cells do originate from circulating blood cells, and circulating chemokines/cytokines are important in activating and recruiting immune cells. Hence, the circulating level of immune cells and cytokine production capacity is probably relevant for local tissue immunity.

We acknowledge that our study has limitations. Firstly, 300DM and 500FG were recruited and measured at two years apart, although established using an identical setting, the same protocol and in the same lab. There may thus be some differences in the absolute immune cell counts or cytokine levels due to a batch effect. Therefore, this study was not designed as a case-control study, but the healthy controls were used to compare genetic associations identified in the T1D cohort. Secondly, young people were overrepresented in 500FG. Although age has been regressed out in both of the cohorts, there might be a bias in the genetic mapping. Finally, our type 1 diabetes-specific analyses should be viewed exploratory as they have not been validated in a separate cohort. Our study has also strengths: it applied cutting-edge technologies to assess immune cell function and genetic variation, and it is the first study which comprehensively combines ‘omics’ technologies to abundant phenotyping in a rather large group of participants to explain between individuals’ variation in immune responses.

## Conclusions

In conclusion, we applied a novel high-throughput functional genomics approach, and show that genetic factors regulate immune responses in T1D patients. We show genetic susceptibility to immune phenotypes in patients with T1D, and highlight the importance of T cell immunity in the genetic regulation of T1D. We identify specific cell populations that are likely involved in pathophysiology of T1D (CCR5+ T-regs). Together, these findings may provide an avenue towards identification of novel preventive and therapeutic treatments.

## Materials and Methods

### Study cohort

This study mainly focuses on a 300DM cohort, and involves a 500FG cohort (part of HFGP*(Netea et al., 2016)*). In total, 132 males and 111 females of Caucasian origin with T1D were recruited in the 300DM cohort at the Radboudumc, the Netherlands. Their age ranges from 20 to 84 years. Detailed information of 500FG cohort can be found in the previous publications(Aguirre-Gamboa et al., 2016; ter Horst et al., 2016; Li et al., 2016).

### Measurement of Immune Cell composition

Myeloid and lymphoid immune cell levels were measured by 10-color flow cytometry. The parental and grandparental proportion from 73 manually annotated immune cells as well as the proportion of CD4+ T cells, CD8+ T cells, memory T regulatory cells and monocytes carrying chemokine receptors: CCR6 (CD196), CXCR3 (CD183), CCR4 (CD194), CCR5 (CD195) and CCR7 (CD197) were calculated respectively, and end up with 269 immune cell traits of the 300DM cohort. We used the same gating strategy of measuring cell subpopulations as 500FG (See reference*(Aguirre-Gamboa et al., 2016)* and Figure 1—figure supplement 2).

### Stimulation of PBMCs and measurement of Cytokine production capacity

Isolation of PBMCs were washed twice with cold phosphate-buffered saline (PBS) and suspended in Roswell Park Memorial Institute (RPMI) 1640 Dutch-modified culture medium (Gibco/Invitrogen, Breda, the Netherlands) supplemented with 50 mg/l gentamycin (Centraform), 1 mM pyruvate (Gibco/Invitrogen) and 2mM L-glutamine (Gibco/Invitrogen). Cells were counted on a Sysmex XN-450 Hematology Analyzer (Sysmex Corporation, Kobe, Japan).

For *in vitro* stimulation experiments, 5x10^5^ cells/well were cultured for 24hr or 7 days at 37°C and 5% CO_2_ in 96-well round-bottom plates (Greiner). For the 7 days’ cultures, medium was supplemented with human pooled serum (pooled from healthy blood donors, end concentration 10%). Supernatants were collected and stored in -20^◦^C until used for ELISA.

The following stimulations were used: (LPS (100ng/ml, 1ng/ml), Pam3cys, *Borrelia* mix, *C. albicans* conidia, *C. albicans* hyphae, Imiquimod (IMQ), *S. aureus*, *S. pneumoniae*, palmitic acid (C16), C16+ monosodium urate crystals (MSU), *E. coli*, *M. tuberculosis* (MTB), *B. burgdorferi*, *C. burnetii*, *C. neoformans*, Oxidized low-density lipoprotein (OxLDL), OxLDL+ LPS, Polyinosinic:polycytidylic acid (Poly IC), *Rhizopus* microspores, *Rhizopusoryzae*)*(Li et al., 2016)*. Concentrations of monocyte-derived cytokines (IL-1β, IL-6, TNF-α, IL-8, IL-10, IL-1Ra, MCP-1) and T cell-derived cytokines (INF-γ, IL-17, IL-22) in response to various stimulations were measured in peripheral blood mononuclear cells (PBMC) by ELISA kits following the manufacturers protocol. For all cytokines commercially available kits were used (R&D Systems, MN, USA or Sanquin, Amsterdam, the Netherlands).

### Genotyping, quality control and imputation

DNA samples of 224 Dutch T1D patients were collected and genotyped by the Infinium® Global Screening Array. The genotype calling was performed using Opticall 0.7.0*(Shah et al., 2012)* with default settings (call rate > 0.99). Two samples were removed due to contamination or mislabeling; one sample from a related individual (identity be descent > 0.185) was removed and six genetic outliers were identified by either heterozygosity rate check (individuals with heterozygosity rate heterozygosity rate ± 3 standard deviations from the mean were excluded) or multi-dimensional scaling (MDS) plots of samples merging with 1000 Genomes data, leaving 215 samples, which show good consistence with European samples from 1000 Genomes data (Figure 1—figure supplement 1) according to standard protocol (Anderson et al., 2010). DNA samples from the 500FG cohort were genotyped by Illumina Human OmniExpress Exome-8 v1.0 SNP chip. Outliers were excluded according to population relationship, medication and disease information. Details can be found in our previous studies in 500FG*(Aguirre-Gamboa et al., 2016; Li et al., 2016)*. Single-nucleotide polymorphisms (SNPs) with a minor allele frequency (MAF) < 0.001 were removed from each cohort, and a Hardy-Weinburg equilibrium (HWE) P value < 1×10^-5^ were removed for the healthy cohort (500FG). By taking shared genetic variants, we merged the 300DM cohort and 500FG cohort, and we used an online genotype imputation service provided by Michigan Imputation Server (https://imputationserver.sph.umich.edu)(Das *et al., 2016)* with HRC Panel 1.1 as reference. SNPs with low imputation quality (R2 < 0.3) and/or MAF < 0.01 in all imputed samples and/or HWE p values < 1×10^-5^ in healthy individuals were excluded leaving 4304387 SNPs and 666 individuals (N_300DM_ = 215 and N_500FG_ = 451).

### Preprocess of the data

Immune cell proportion was calculated by dividing counts of their parental and grandparental cell types (Supplementary file 6). An inverse rank transformation(Aguirre-Gamboa et al., 2016; Orrù et al., 2013) was applied on the proportion values for genetic association analysis. Cytokine levels were log2 transformed.

### Immune parameter QTL mapping

After intersecting with available genotype data and excluding volunteers with a mixture or another genetic background, 214 were left for immune parameter quantitative trait locus (QTL) mapping. We next evaluated covariates influencing immune function. Associations with age, gender, seasonal effects were calculated using previously described methods(Aguirre-Gamboa et al., 2016; ter Horst et al., 2016; Li et al., 2016), i.e. Spearman correlation analysis and linear regression model. Significance was declared after multiple testing correction (FDR < 0.05). Age significantly associated with 129/269 immune cell traits and 12/55 cytokine traits; gender associated with 59/269 immune cell traits and 5/55 cytokine traits; seasonal effects associated with 121/269 immune cell traits and 38/55 cytokine traits (Figure 4—figure supplement 3). Thus, we took age, gender and seasonal effects as covariates in the linear regression for both immune cell proportion QTL mapping as well as cytokine QTL mapping. Besides, considering the effect of immune cell proportion on cytokine production, we took major cell types, including monocyte, lymphocyte, T cell, B cell and NK cell proportion in PBMC as covariates in a linear model for cytokine QTL mapping, as we previously did*(Li et al., 2016)*. Linear regression function was applied with an R package Matrix-eQTL*(Shabalin, 2012)* for immune parameter QTL mapping, and software METAL*(Willer et al., 2010)* was applied to summary statistics from both cohorts for meta-analysis, in which the model based on effect size and standard error with default settings were used. P values < 5×10^-8^ were considered to be genome-wide significant. Lambda values were calculated indicating no obvious inflations (0.977-1.026).

### Extraction of T1D GWAS SNP list

We downloaded a summary of T1D GWAS results from GWAS-catalog (https://www.ebi.ac.uk/gwas/)(Buniello *et al., 2019)* in November 2019, and removed studies done in non-European-ancestry populations. Top SNPs within LD (r^2^ > 0.1) from different studies were considered as the same locus, and SNPs with the lowest p value were taken into analysis. We noticed that the effect directions of some SNPs were unclear or inconsistent in different studies. In this case, we assigned directions according to the newest GWAS study*(Onengut-Gumuscu et al., 2015)*.

### GWAS analysis of 300DM cohort for the known T1D loci

We extracted all proxies in strong LD with top SNPs from published T1D GWAS (Case-Control) studies (r^2^ > 0.8) and performed a chi-square test on clinical status by using PLINK 1.9. Samples in 300DM were taken as cases and samples in 500FG were controls.

### Impact of T1D GWAS loci on immune phenotypes

In order to detect the impact of T1D GWAS loci on immune cell populations, we grouped all traits into four categories (B cells, T cells, monocytes and NK cells), and counted the number of suggestive associations (P value < 0.05) between 63 top SNPs from T1D GWAS loci and immune cell traits. 1000 permuted sets of 63 SNPs were randomly selected from independent SNPs (r^2^ > 0.2) pruned from all genotyped SNPs. We then compared the associations of the 63 top GWAS SNPs with associations between 1000 permutated sets and the same category of immune traits. P value was calculated by the percent of association numbers from permuted sets greater than the association number of the 63 T1D GWAS SNPs.

We further applied a multivariate linear model to estimate the proportion of variance of each immune phenotype explained by the top SNPs from T1D GWAS loci. We repeated this analysis on 1000 permuted sets of 63 independent SNPs, which were used as reference set. We then compared the null distribution with variance explained by 63 top SNPs from T1D GWAS loci. P value was calculated by the percent of explained variance from permuted sets larger than the variance explained by the 63 T1D GWAS SNPs.

### Gene expression analysis on *CCR5* and its corresponding ligands genes

Normalized gene expression data in PBMCs and Pancreases from T1D and controls were acquired from https://www.ebi.ac.uk/gxa(Papatheodorou *et al., 2019)*. A *Student’s* t-test was applied to compare gene expression between groups.

### Post-QTL analysis

An R package ggplot2 was used to generate Manhattan plots and boxplots. Locus-zoom plots were made by an online tool(http://locuszoom.org)(Pruim *et al., 2010)*. We used an R package coloc*(Giambartolomei et al., 2014)* to perform colocalization analysis with T1D GWAS summary statistics and immune parameter QTLs profile. For pathway analysis, genes located within +-10kb of genome wide significant SNPs (p value < 5×10^-8^) were extracted and analyzed by an online tool FUMA*(Watanabe et al., 2017)* (https://fuma.ctglab.nl).

ANNOVAR was used for annotating genetic variants*(Wang et al., 2010)*.

### Other packages used in this paper

Pheatmap was used to make heatmaps. Scatter plots and bar plots were generated by ggplot2.

## Data Availability

Results reported in the manuscript has been summarised in supplementary tables. Codes for generating all results could be found at github (https://github.com/Chuxj/Gf_of_ip_in_T1D). Raw immune phenotype data (cell proportion and cytokine production in 300DM) and summary statistics could be found at Dryad (https://doi.org/10.5061/dryad.4f4qrfjd0). Genetics and donor information that could compromise research participant privacy are only vailable upon request to the corresponding authors (http://hfgp.bbmri.nl).

https://github.com/Chuxj/Gf_of_ip_in_T1D

https://doi.org/10.5061/dryad.4f4qrfjd0

## Acknowledgements

We thank all of the volunteers in the 300DM and 500FG for their participation. We thank Marc J. Bonder for discussions.

## Author contributions

Y.L., C.T. and M.G.N designed the study. X.C. performed statistical analysis supervised by Y.L., A.J., H.K., X.H. and I.J. performed the experiments and processed the data. L.C. and C.X. helped with the data analysis. Y.L., C.T., M.G.N, C.W and X.C interpreted the data. Y.L., C.T., M.G.N. and X.C wrote the manuscript with input from all authors.

## Data and code availability

All raw data of immune phenotypes have been deposited in Dryad (https://doi.org/10.5061/dryad.4f4qrfjd0). Individual genetic data and other privacy-sensitive individual information are available upon request to corresponding authors by email or at Human Functional Genomics Project website (http://hfgp.bbmri.nl). This dataset is not publicly available because it contains information that could compromise research participant privacy, while summary statistics directly generated from genetic data that will precisely reproduce published results are all deposited in Dryad (https://doi.org/10.5061/dryad.4f4qrfjd0). Custom scripts for generating summary statistics and all results are deposited in GitHub (https://github.com/Chuxj/Gf_of_ip_in_T1D).

## Ethical statement

The 500FG-DM study was approved by the ethical committee of Radboud University Nijmegen (NL-number: 54214.091.15). Experiments were conducted according to the principles expressed in the Declaration of Helsinki. Written informed consent was obtained from all participants.

## Figure supplement legends

**Figure 1 - figure supplement 1.**
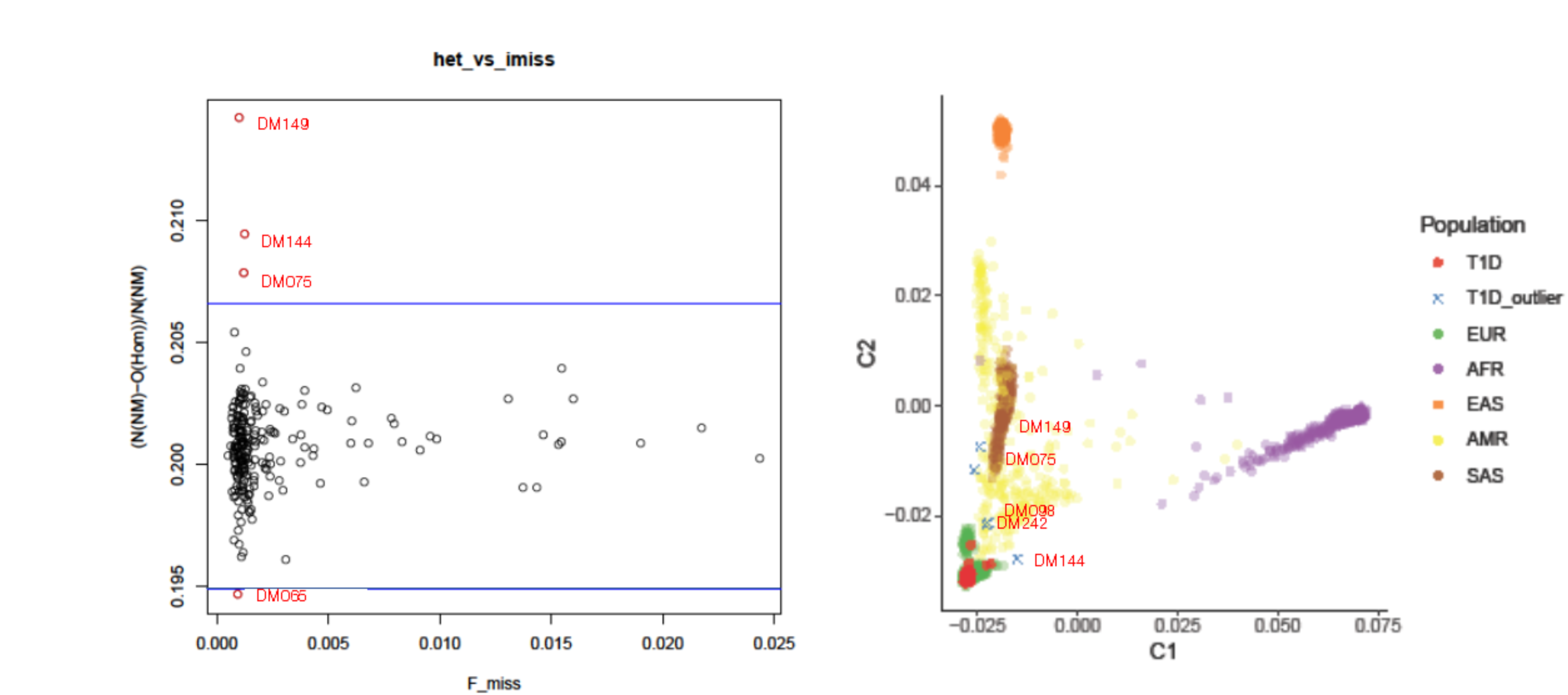
Heterozygosity check. Left-side panel, an overview of missingness vs heterozygosity for all samples represented by circle; x-axis: Proportion of missing genotypes, y-axis: Heterozygosity rate) and ancestry clustering. Right-side panel, multiple dimensional scaling plot for our T1D samples (red dots) in comparison with reference samples from 1000G. X and y axis correspond to component 1 and component 2 in multiple dimensional scaling plot, respectively. In total, six outliers (red symbols from either left or right panel) were removed in further analysis.

**Figure 1 - Figure supplement 2.**
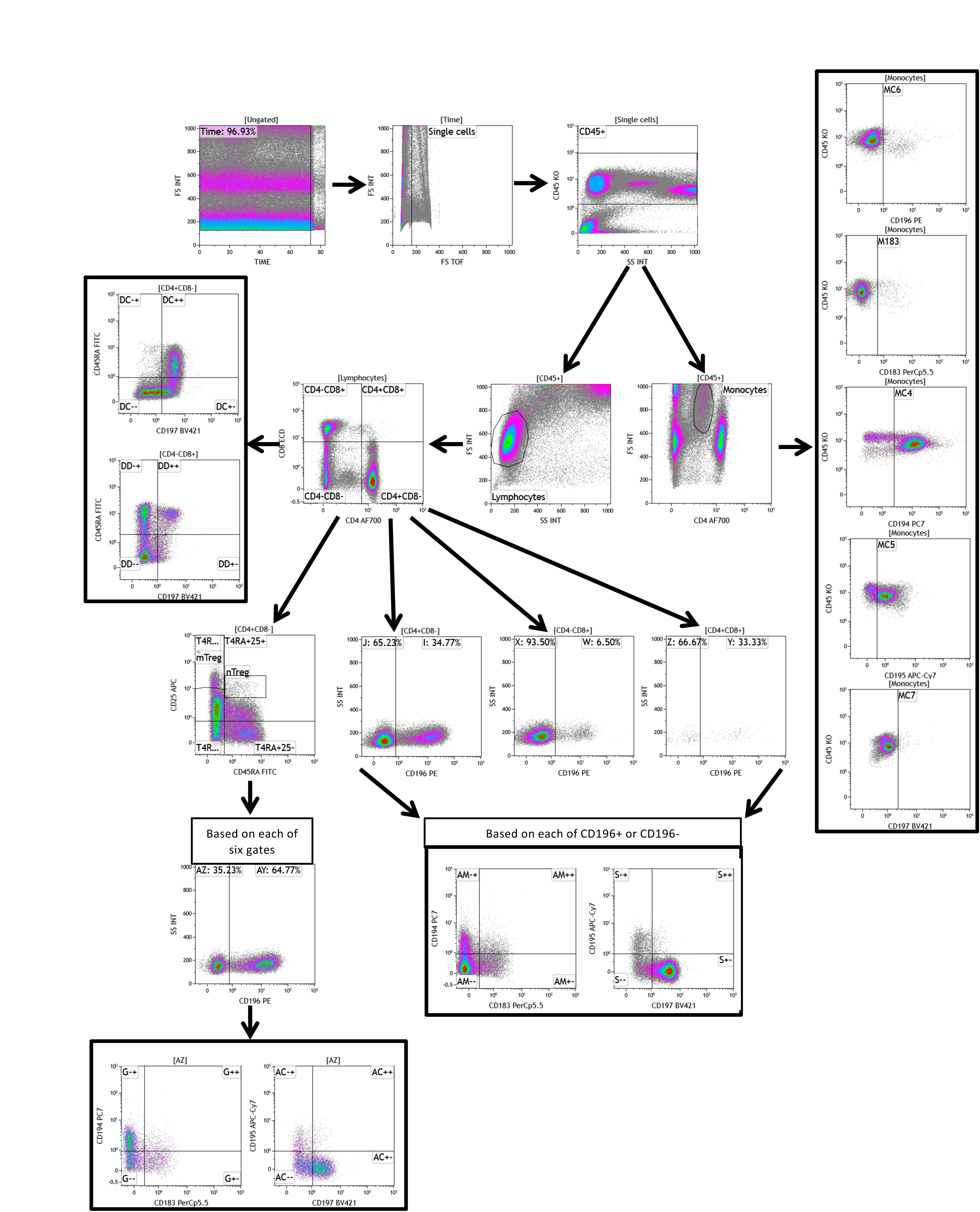
Flow cytometry gating strategy of the chemokine receptor panel.

**Figure1 - Figure supplement 3.**
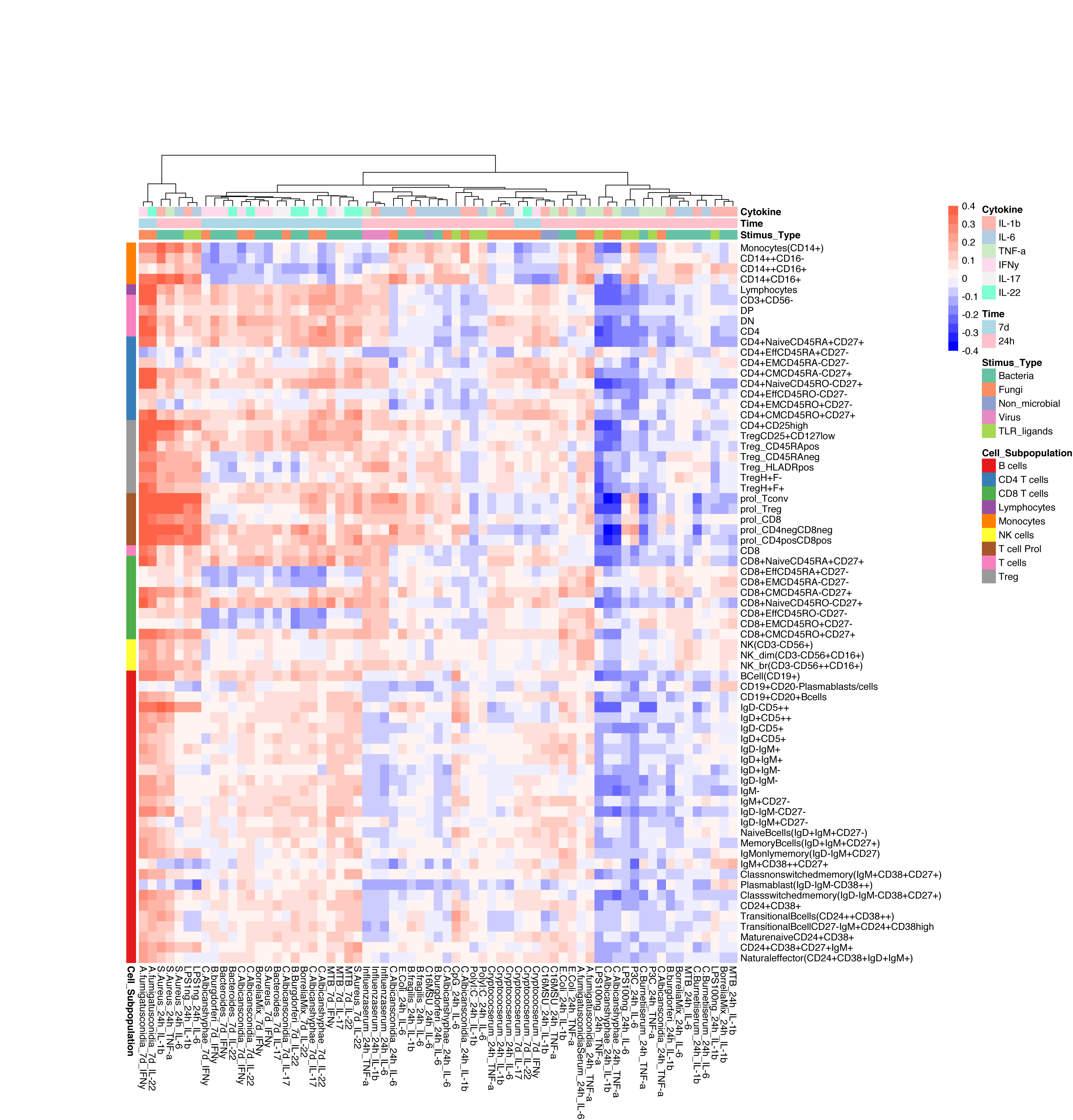
Correlation between cell counts and cytokine production in healthy individuals. Heatmap of correlation coefficients (indicated by colors) between immune cell counts (y-axis) and cytokine production in response to stimulations (x-axis) in healthy individuals (500FG), which is consistent with 300DM.

**Figure 2 - Figure supplement 1.**
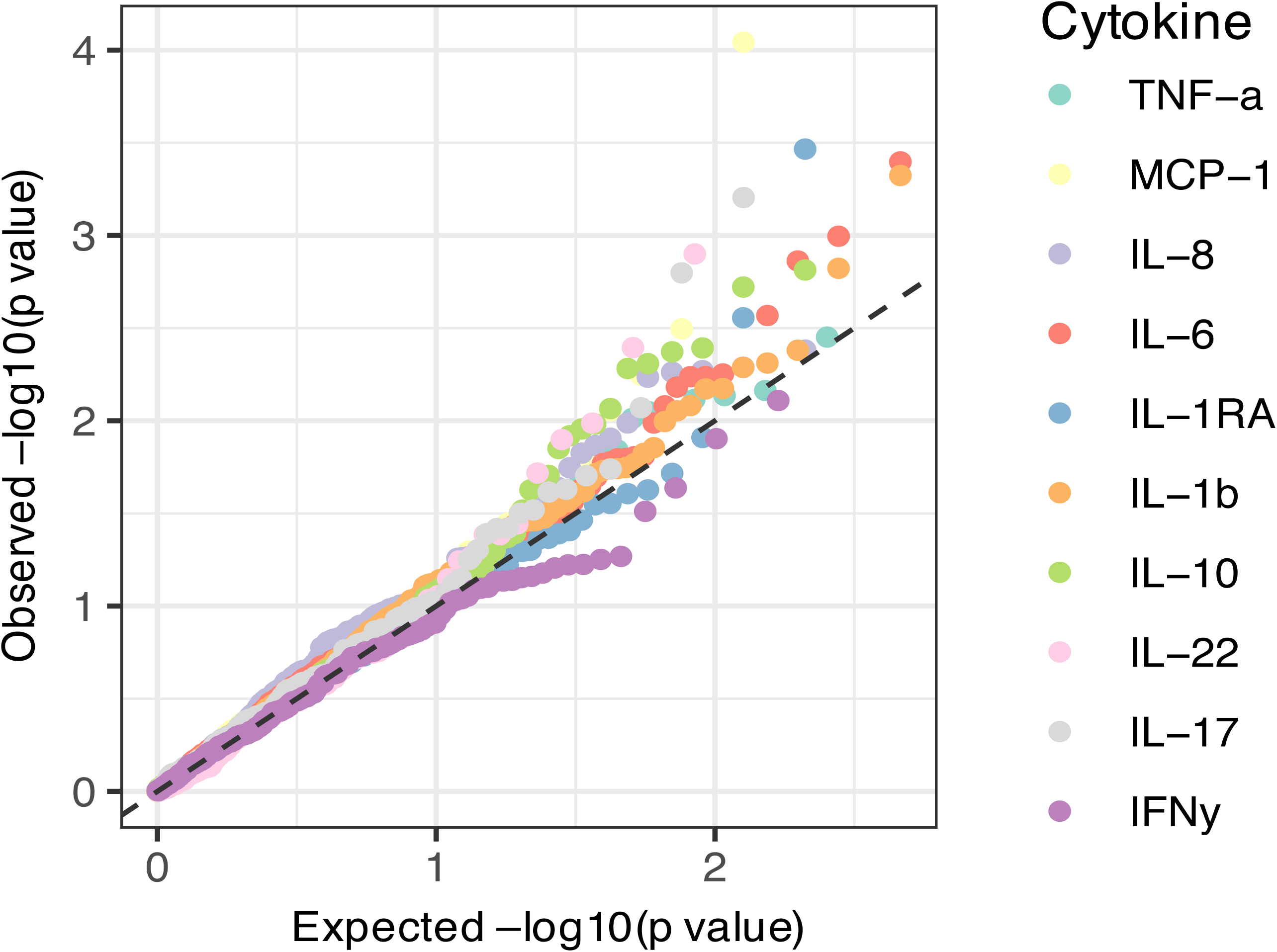
QQplots of QTL profiles of 62 T1D GWAS loci grouped by cytokine types.

**Figure 2 - Figure supplement 2.**
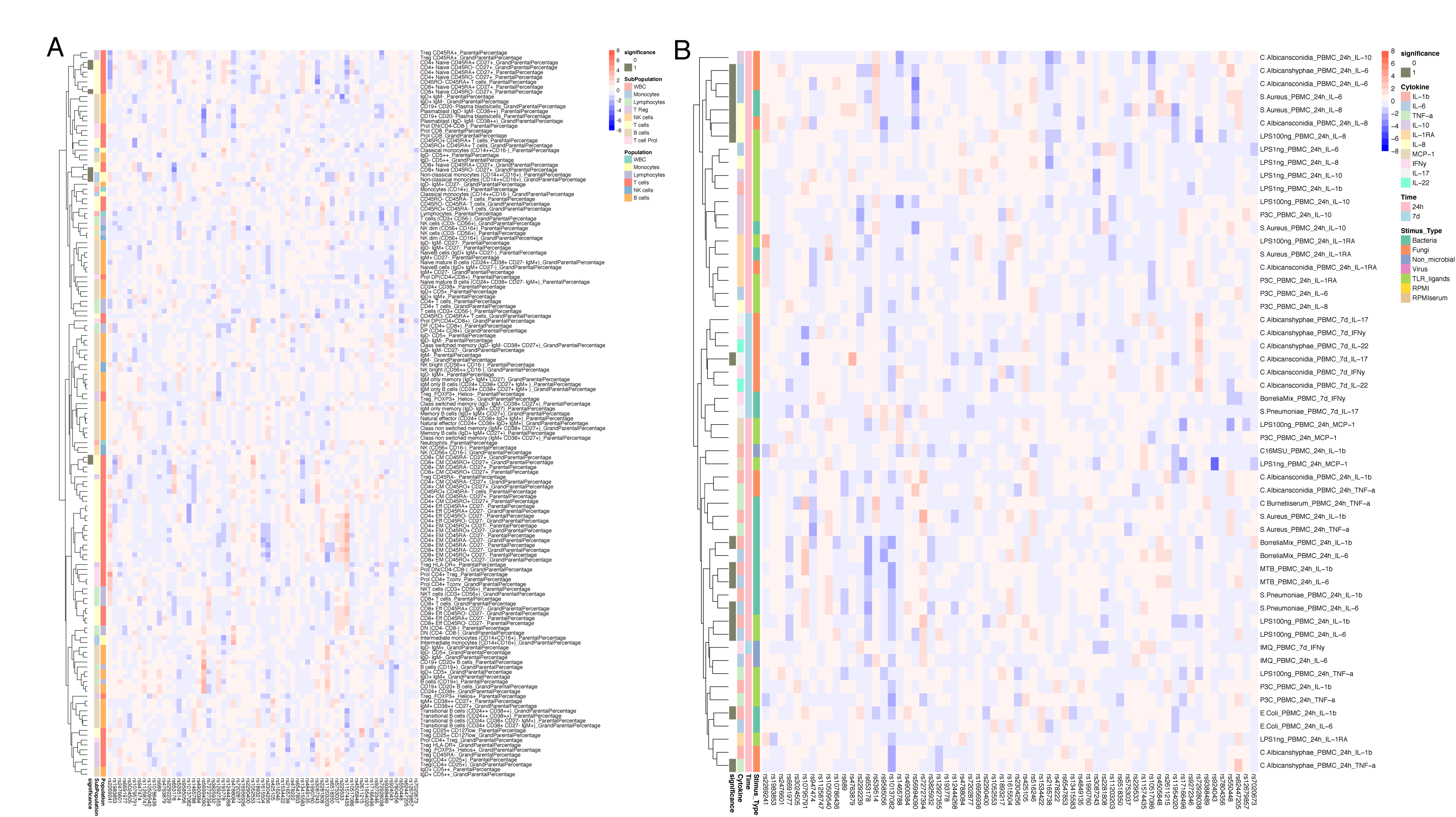
Impact of T1D GWAS SNPs on immune phenotypes. **A**,**B**, Heatmap of QTL profiles of immune phenotypes across 62 T1D GWAS loci, colored by - log10(p values) and effect direction of T1D risk allele (**A**: immune cell proportion traits; **B**: cytokine production upon stimulations).

**Figure 4 - Figure supplement 1.**
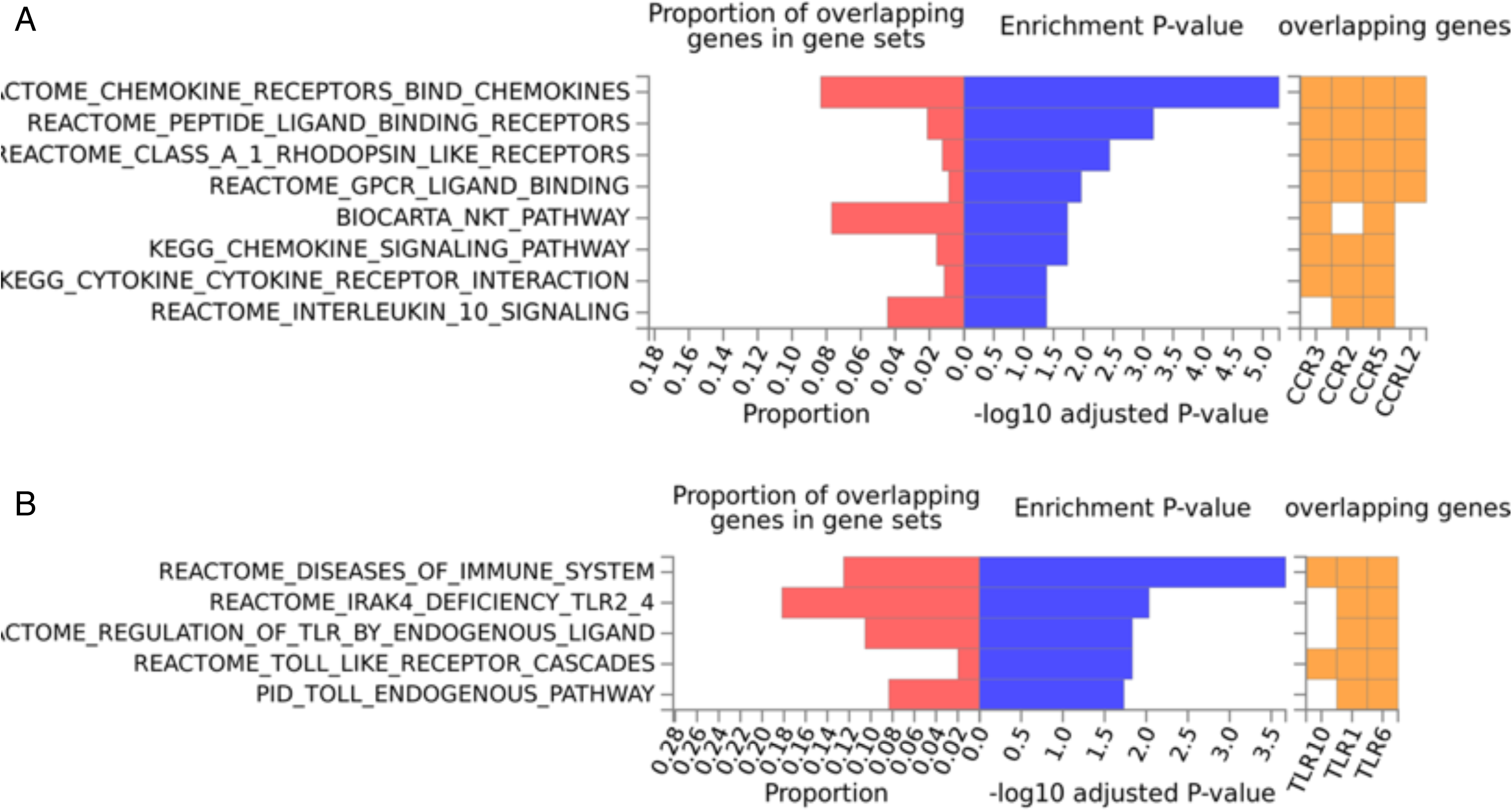
Enriched pathways of QTLs found for immune phenotypes. Pathway enrichment analysis using genes located within immune cell proportion QTLs (**A**) and cytokine production QTLs (**B**).

**Figure 4—figure supplement 2.**
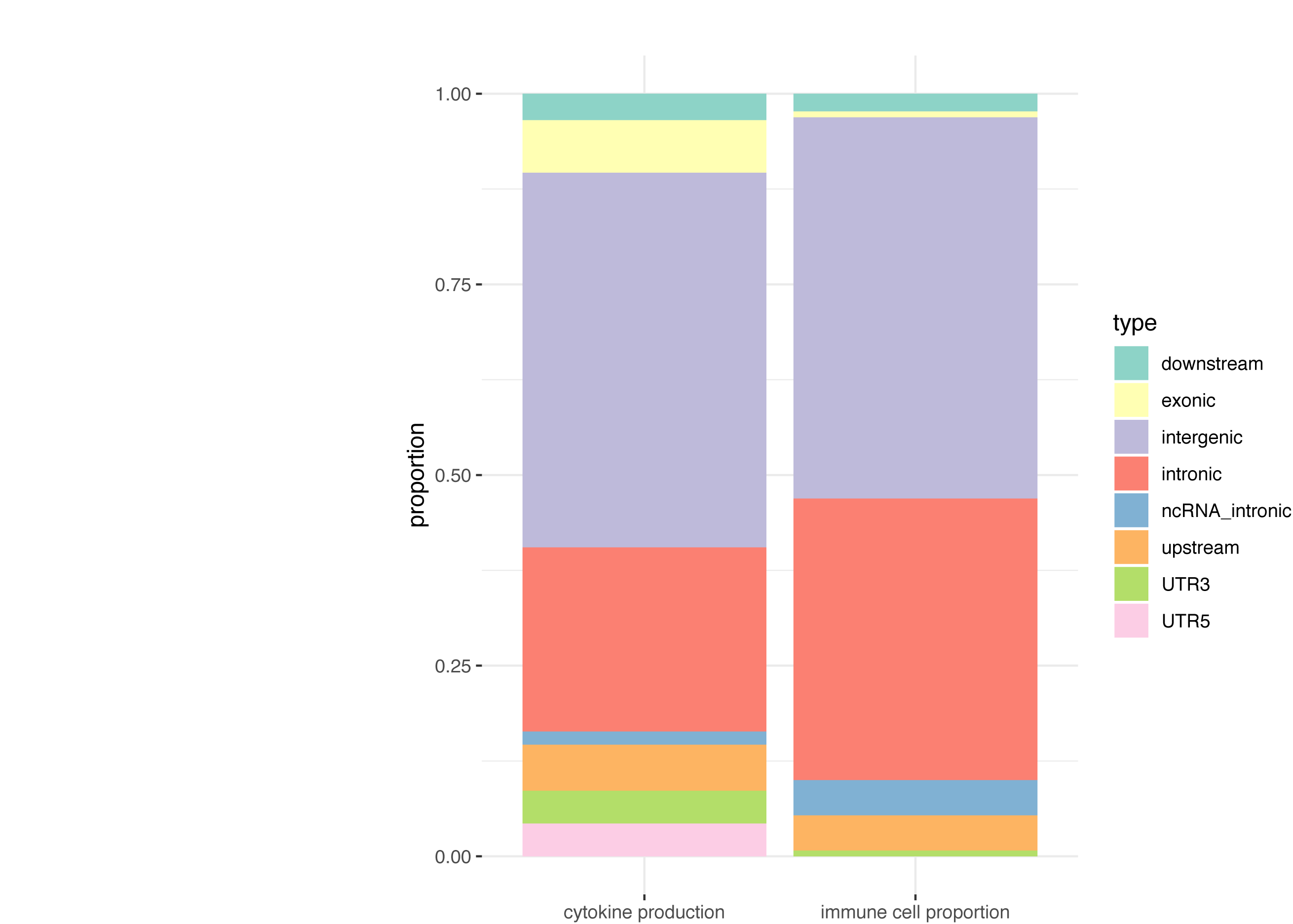
Functional annotation of the immune phenotypes associated SNPs (P < 5×10^-8^). Stacked boxplots showing the percentage of genomic elements where the immune phenotypes associated SNPs located.

**Figure 4—figure supplement 3.**
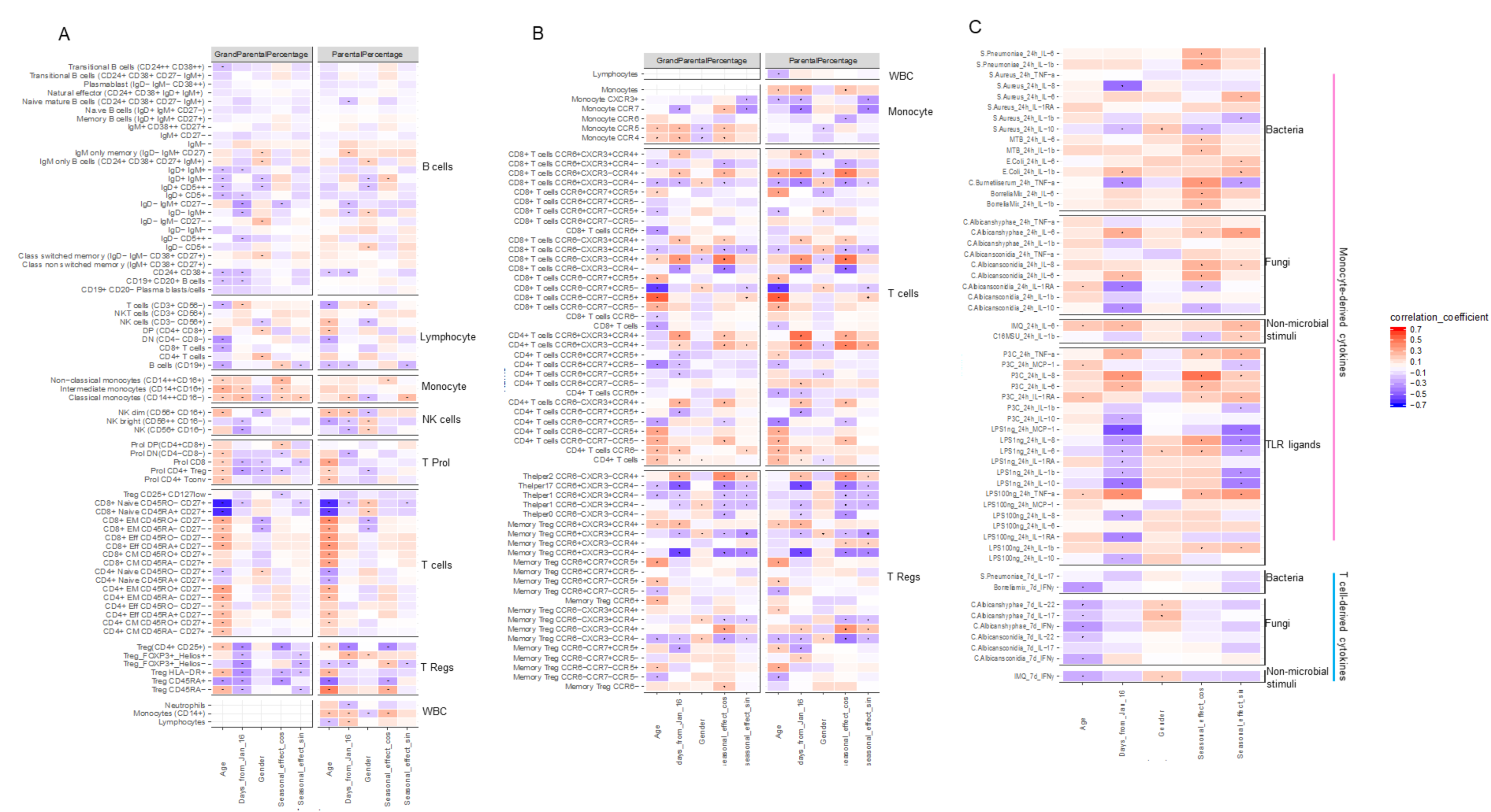
Impact of age, gender and seasons on immune phenotypes. **A, B**, Heatmaps of correlation between immune cell counts (y-axis) and age, gender, seasons (x-axis). Correlation coefficients were indicated by colors, and significant correlations (FDR <0.05) are labeled by dots. **C**, Heatmap of correlation between cytokines levels upon stimulations and age, gender, seasons. Correlation coefficients were indicated by colors, and significant correlations (FDR <0.05) are labeled by dots.

